# Liver functional radiomics of gadoxetic acid-enhanced magnetic resonance imaging: A proof-of-concept study

**DOI:** 10.64898/2026.09.09.26362661

**Authors:** Qiang Wang, Aristeidis Grigoriadis, Stefan Gilg, Antonios Tzortzakakis, Ernesto Sparrelid, Torkel B. Brismar

## Abstract

**Objectives:** This study aimed to identify reproducible and repeatable radiomics features (RF) of gadoxetic acid-enhanced MRI that correlate with liver function.

**Methods:** This study included 10 patients with colorectal liver metastasis who received a right hepatectomy and were prospectively recruited. Before surgery, all patients underwent indocyanine green (ICG) retention test, hepatobiliary scintigraphy (HBS) exam and gadoxetic acid-enhanced MRI. RF were extracted from the non-tumoral liver parenchyma at the MRI images before contrast injection and the hepatobiliary phase. Delta-RF were calculated by RFs of hepatobiliary phase subtract those before contrast injection. ICG-R15, HBS results, and kinetic growth rate (KGR) of the future liver remnant volume were used as quantitative indices of liver function. Reproducible and repeatable RF were identified via inter-and intraclass correlation analysis with a coefficient threshold of 0.80. Correlations between RF/delta-RF and the three liver function indices (ICG-R15, HBS and KGR) were evaluated by Spearman correlation analysis.

**Results:** One hundred and seven RFs were extracted from each MRI phase (214 in total and 107 derived delta RFs). With inter-and intraclass correlation analysis, 60 reproducible and repeatable RFs were identified. After removing RFs with higher correlation coefficients, 12 RF from images before contrast injection, 23 RF from hepatobiliary phase, 29 delta-RF were identified significantly correlated with the liver function metrics (ICG-R15, HBS and KGR).

**Conclusions:** A set of reproducible and repeatable radiomics features of gadoxetic acid-enhanced MRI which were associated with liver function were identified. These features can be adopted for future liver function-related radiomics research.

## Introduction

Accurate liver function assessment before surgery is of clinical importance to ensure safe liver resection and low risk of post-hepatectomy liver failure in the treatment of liver tumors [1].

Traditionally, clinical scoring systems such as Child-Pugh score, Albumin-Bilirubin grade, and quantitative approaches like indocyanine green (ICG) retention test are used for liver function evaluation [2]. However, these methods only provide global information, failing to capture the heterogeneous distribution of liver function across the liver segments, which is a critical consideration for pre-operative planning, especially in major hepatectomies with insufficient future liver remnant volume [3].

Gadoxetic acid-enhanced magnetic resonance imaging (MRI), along with ^99m^Tc mebrofenin hepatobiliary scintigraphy (HBS), offers a promising alternative to provide regional liver function information [4]. Gadoxetic acid and ^99m^Tc mebrofenin are both actively taken up by hepatocytes via organic anion-transporting polypeptides transport mechanisms [5]. While HBS has been a reference for regional liver function evaluation, it is hampered by lower spatial resolution, radiation exposure and prolonged tracer uptake time [6]. In contrast, gadoxetic acid-enhanced MRI has emerged as a valuable tool for quantitatively evaluating liver function over the past decade [7]. Previous studies have demonstrated a solid correlation between parameters derived from gadoxetic acid-enhanced MRI (such as the liver-to-muscle ratio of signal intensity) and established measures of liver function, such as the ICG retention rate at 15 minutes (ICG-R15), Child-Pugh score, and Model for End-Stage Liver Disease score [8, 9]. This suggests its potential to provide a more comprehensive pre-operative assessment that integrates morphological, volumetric, and functional capacities.

In recent years, radiomics have drawn extensive attention in the biomedical field due to its powerful predictive efficacy[10, 11]. Radiomics can extract a large number of quantitative imaging features from daily used medical images such as MRI and be incorporated into prediction models for improvement of diagnosis and prognostication [10]. It provides valuable insights into imaging phenotype and tissue/tumor pathophysiologic behaviors. Previous studies have applied radiomics of gadoxetic acid-enhanced MRI for predicting postoperative liver failure [12–14]. However, a direct investigation on the relationship between the radiomics features (RFs) and the established liver function indices has not been performed.

Understanding the relationship between RF and liver function indices could offer valuable insights into the underlying biological meaning of imaging features, provide a reliable tool for regional liver function assessment, and ultimately lead to an important part of personalized treatment strategies. Therefore, this study aims to identify reproducible liver function associated RFs in a well-designed, prospective cohort where patients with colorectal liver metastasis (CRLM) underwent ICG test, HBS exam and gadoxetic acid-enhanced MRI at several predefined time-points during the perioperative period [15].

### Patients and methods Study design and subjects

This study is a secondary analysis of our previous study in which we prospectively recruited 10 CRLM patients who underwent ICG test, HBS exam and gadoxetic acid-enhanced MRI sequentially before surgery (consisting of a right hepatectomy in all patients), and on postoperative day 7 and day 28 at Karolinska University Hospital [15]. In the present study, the data before surgery were used, i.e. ICG retention rate at 15 minutes (ICG-R15), HBS examinations (the result of the total liver functional volume corrected by body surface area and expressed as uptake rate of mebrofenin, %/min/m^2^, was used), and gadoxetic acid-enhanced MRI images before intravenous contrast media injection and at hepatobiliary phase. In addition, liver kinetic growth rate (KGR) was obtained by calculating the degree of hypertrophy (future liver remnant volume after hepatectomy on postoperative day 7 subtracted by future liver remnant before surgery, then divided by one week and expressed as %/week). The processes and methods of the ICG test, HBS exam and liver volume calculation have been previously described [15]. The study workflow is shown in Figure 1. The research protocol was approved by the Regional Ethical Review Board (Dnr 2012/583-31/4) and informed consent was obtained from all participating patients.

**Figure 1.**
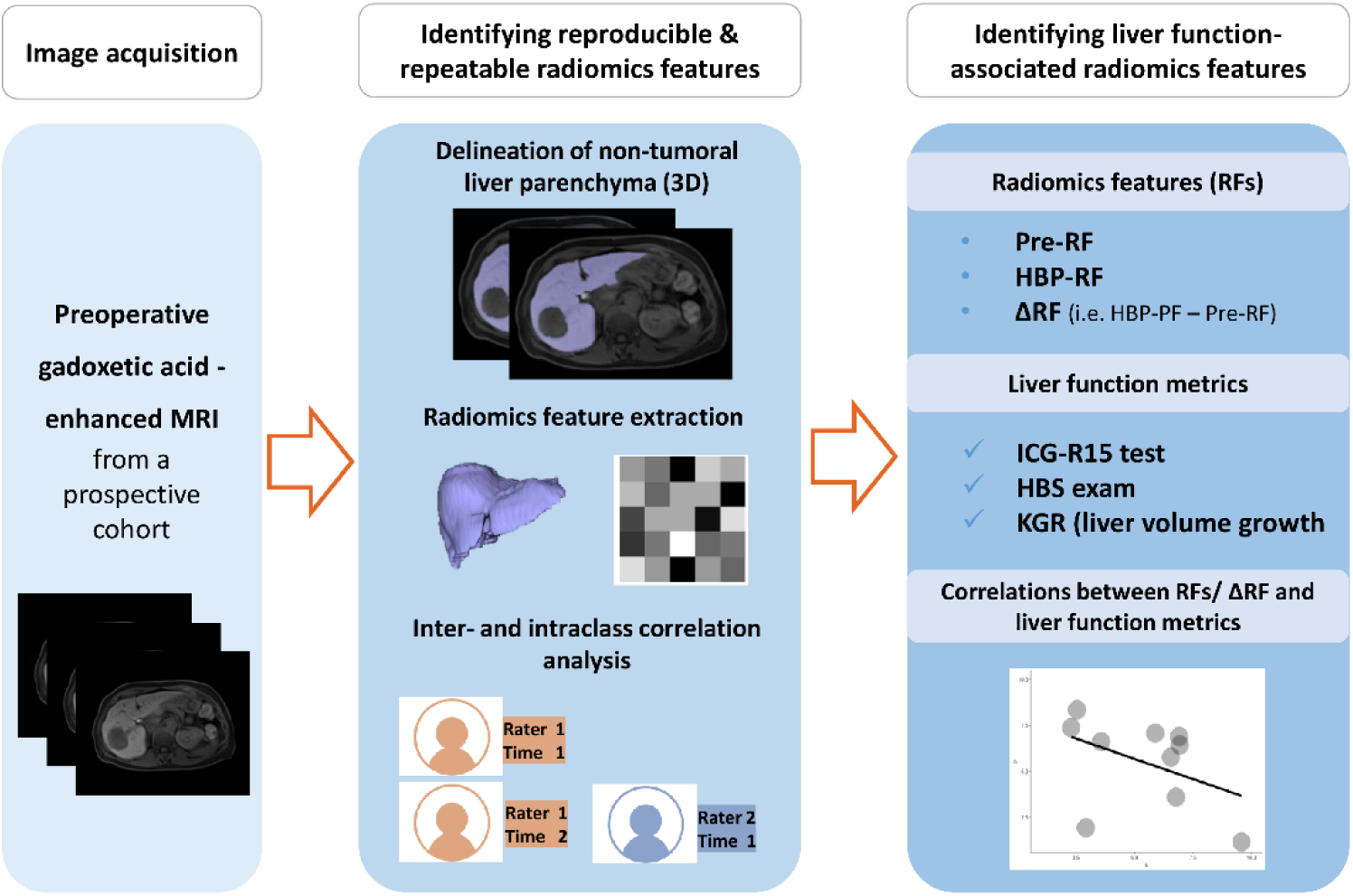
Study design and workflow.

### Gadoxetic acid-enhanced MRI exam

MRI examination was conducted using a 1.5 T MRI scanner (Aera, Siemens Healthineers, Erlangen, Germany) equipped with a four-channel sensitivity encoding body coil. Dynamic contrast-enhanced images were obtained utilizing a T1 weighted fat suppressed three-dimensional volume interpolated breath-hold examination sequence, with the following scanning parameters: repetition time of 4.1 ms, echo time of 1.95 ms, flip angle of 10°, field of view of 415 mm, acquisition matrix resolution of 256 × 156, 48 slices, and a slice thickness of 5 mm. Gadoxetic acid (Primovist®, Bayer Healthcare, Berlin, Germany) was administered intravenously via the anterior cubital vein at a rate of 2 ml/s using a power injector, with a concentration of 0.25 mmol/ml and a dosage of 0.1 ml/kg body weight. Hepatobiliary phase (HBP) was obtained at 20 min after gadoxetic acid injection.

In this study, T1WI before and 20 min after contrast media injection (short for “Pre”, “HBP”, respectively) were chosen for radiomics analysis as they are commonly used in signal intensity-based quantification of liver function [7].

### Segmentation of non-tumoral liver parenchyma

Delineation of the non-tumoral liver parenchyma at hepatobiliary phase gadoxetic acid-enhanced MRI was manually performed by two researchers (Q.W & A.G, with abdominal imaging experience of 5 and 12 years, respectively) independently using the software ITK-SNAP (version 3.6.0) in a slice-by-slice manner. One month later, one researcher (Q.W) conducted the same operation again to delineate the non-tumoral liver parenchyma; additional delineation of the non-tumoral liver parenchyma at Pre-MRI images was also performed at this time, and the delta-RF was calculated by (RF at HBP – RF at Pre-MRI images).

### Radiomics feature extraction

Prior to the features extraction, images underwent resampling to achieve a voxel dimension of 1 × 1 × 1 mm³, with the width of the intensity histogram bins set to 25. The Python package “PyRadiomics” (version: 3.0.1, available at https://github.com/AIM-Harvard/pyradiomics) was used to extract the features from the segmented non-tumoral liver parenchyma part. The extracted features fell into the following seven categories: (1) Shape features in both 2D and 3D (14 features), (2) First-order statistical features (18 features), (3) Features derived from the Gray Level Co-occurrence Matrix (24 features), (4) Features derived from the Gray Level Run Length Matrix (16 features), (5) Features derived from the Gray Level Size Zone Matrix (16 features), (6) Features derived from the Gray Level Dependence Matrix (14 features), and (7) Features from the Neighboring Gray Tone Difference Matrix (5 features), leading to a comprehensive extraction of 107 radiomics features in total. The feature set extracted through “PyRadiomics” aligned with the Image Biomarker Standardization Initiative standards [16].

### Identification of reproducible and repeatable RFs

As RF extraction mainly depends on manual segmentation of the volume of interest, inter-and intraclass correlation analyses were performed to evaluate the reproducibility and repeatability of RFs [17]. Interclass correlation coefficients of RF were calculated on the HBP results from the two researchers, and intraclass correlation was performed using the two times of HBP results from the same researcher (Q.W). RFs with inter-and intraclass correlation coefficients > 0.80 were regarded as reproducible and repeatable features. These identified RFs were applied to the Pre-MRI derived RFs (short for “Pre-RF”). Delta-RFs were calculated using these reproducible and repeatable RFs.

### Identification of liver functional RFs

Considering there is an inherent correlation among the large number of RFs when performing replicative first-order and second-order statistics on the images [18], correlation analyses between any two RFs/delta-RFs within each set were performed. One would be randomly abandoned if their coefficient was > 0.99, while the other was kept. Next, to identify liver function associated RFs/delta-RFs, the correlation between these features and liver function indices (represented by ICG-R15, HBS test, and KGR) was evaluated by Spearman’s rank correlation analysis.

### Statistical analysis

All statistical analysis and data visualization were performed in R software (R Foundation for Statistical Computing, Vienna, Austria). An R package “irr” was used to perform inter/intraclass correlation analysis (for interclass: two-way random-effects model, absolute agreement, and single measurement; for intraclass: two-way mixed-effects model, absolute agreement, and single measurement). A p < 0.05 was regarded as statistically significant.

## Results

The basic characteristics of the 10 CRLM patients are shown in Table 1.

**Table 1.**
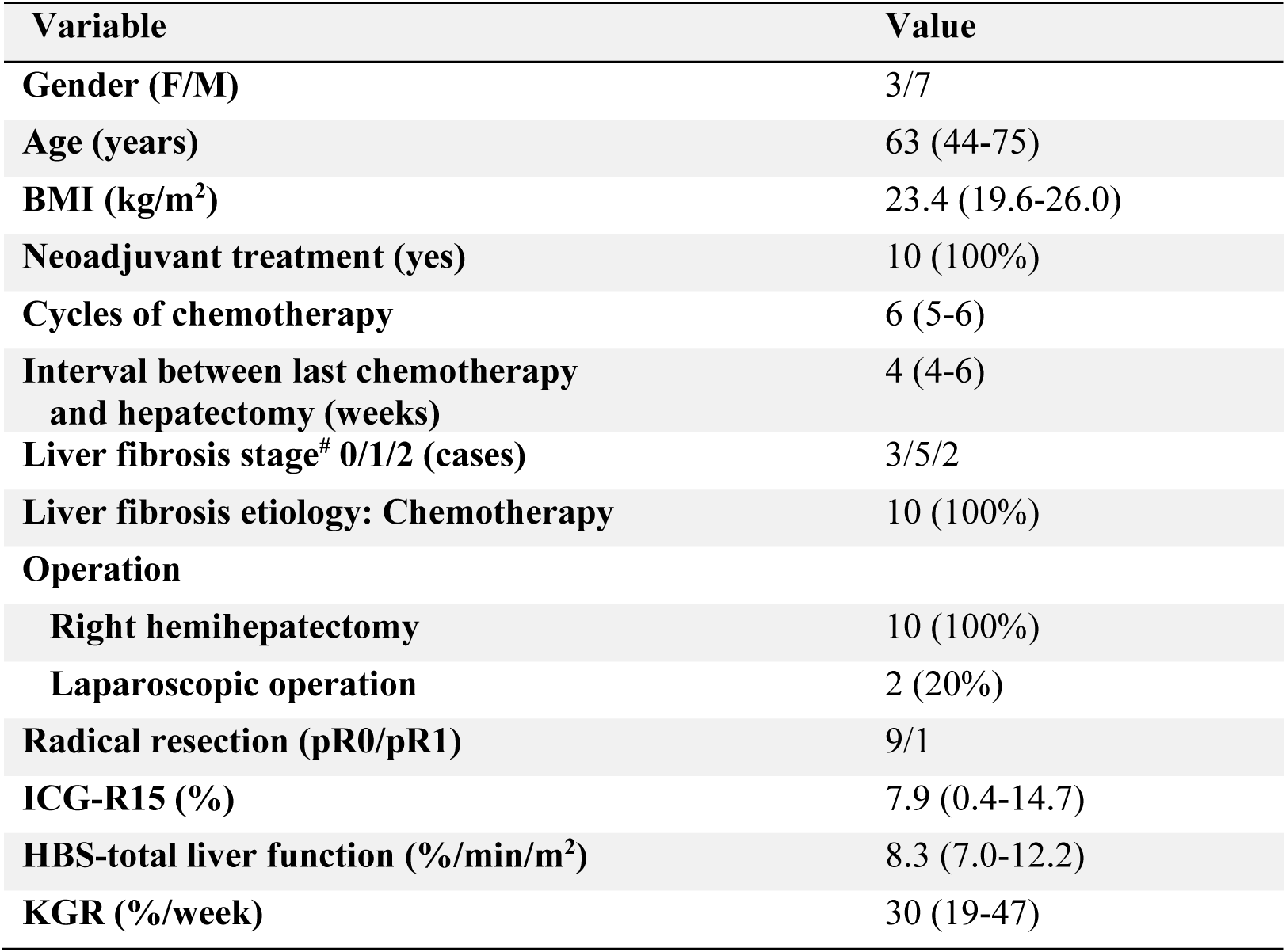
Basic characteristics of the participants.

| Variable | Value |
| --- | --- |
| Gender (F/M) | 3/7 |
| Age (years) | 63 (44-75) |
| BMI (kg/m<sup>2</sup>) | 23.4 (19.6-26.0) |
| Neoadjuvant treatment (yes) | 10 (100%) |
| Cycles of chemotherapy | 6 (5-6) |
| Interval between last chemotherapy and hepatectomy (weeks) | 4 (4-6) |
| Liver fibrosis stage<sup>#</sup> 0/1/2 (cases) | 3/5/2 |
| Liver fibrosis etiology: Chemotherapy | 10 (100%) |
| Operation |  |
| Right hemihepatectomy | 10 (100%) |
| Laparoscopic operation | 2 (20%) |
| Radical resection (pR0/pR1) | 9/1 |
| ICG-R15 (%) | 7.9 (0.4-14.7) |
| HBS-total liver function (%/min/m<sup>2</sup>) | 8.3 (7.0-12.2) |
| KGR (%/week) | 30 (19-47) |

*Note: continuous data are presented as median with range, and categorical data as numbers with percentages. # by the Batts-Ludwig scoring system, in which stage 0, 1 and 2 indicate no, mild, and moderate fibrosis respectively (on a 5-graded scale); BMI, body mass index; pR0, > 1 mm from tumor to resection margin; pR1, < 1 mm from tumor to resection margin.*

### Reproducible and repeatable RFs

One hundred and seven RFs were extracted from the Pre-and HBP MRI images, respectively. In the intraclass correlation analysis, 74 RFs remained after removing the ones with correlation coefficients less than 0.80. Further interclass correlation analysis identified 61 RFs with correlation coefficients greater than 0.80. The number of overlapping RFs in both analyses was 60 (Figure 2A). The distribution of the categories of these reproducible and repeatable RFs is described in Figure 2B.

**Figure 2.**
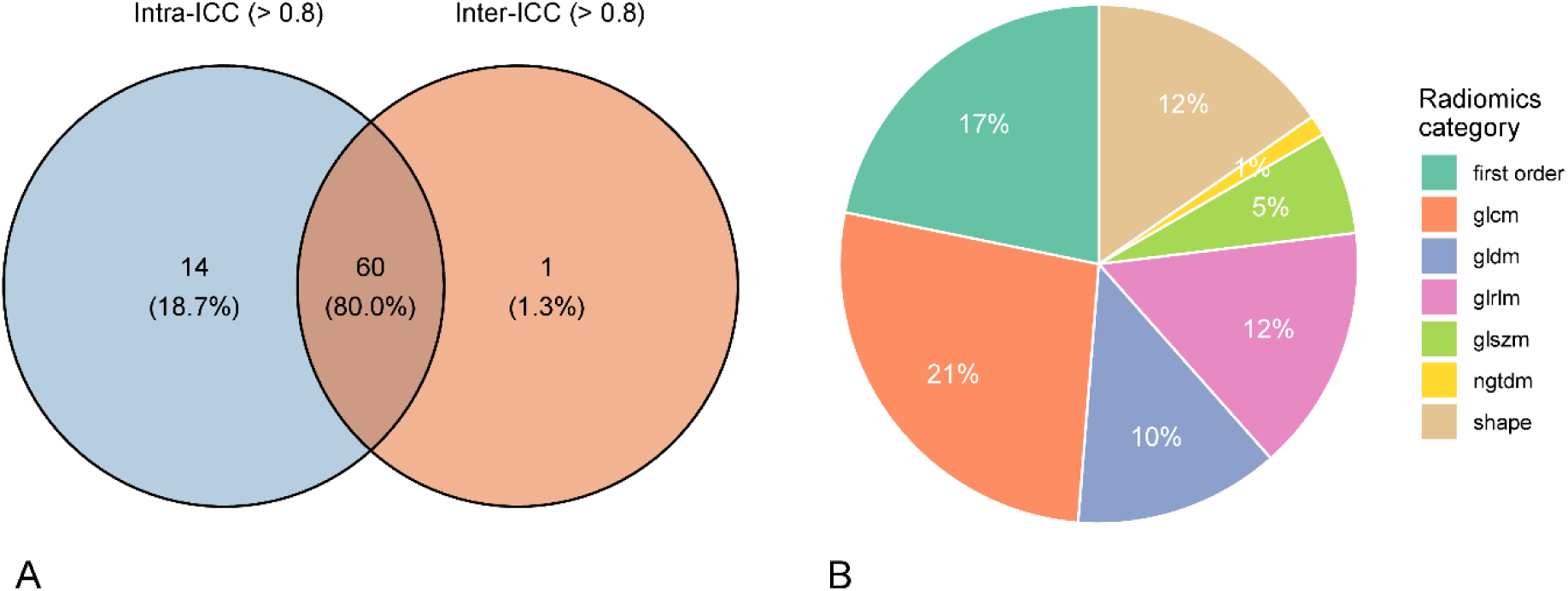
Selection and distribution of reproducible and repeatable radiomics features. **A**. Venn diagram shows radiomics features with high inter-and intraclass correlation coefficients (> 0.80). **B**. Distribution of the reproducible and repeatable radiomics features from hepatobiliary phase gadoxetic acid-enhanced MRI.

The same set of reproducible and repeatable RFs were applied to unenhanced MRI images to obtain the same number of Pre-RFs, and delta-RFs were calculated accordingly by HBP-RFs subtracted by Pre-RFs (n = 60). Correlation analyses between any two RFs/delta-RFs within each RF set were performed to remove “redundant” RFs, and 42 Pre-RFs, 39 HBP-RFs, and 49 delta-RFs were kept after randomly removing one of the RFs with a correlation coefficient > 0.99 in a correlation analysis pair (Supplementary Figure S1).

### Liver functional RF

Further correlation analyses were performed to detect RFs closely correlated with liver function/growth. A total of 7 Pre-RFs, 18 HBP-RFs, and 20 delta-RFs were significantly associated with at least one of the three liver function metrics (i.e. the ICG-R15 test, HBS exam or KGR), corresponding to 12, 23, 29 significant correlation pairs, respectively (Figure 3, Supplementary Figure S2). The full list of these RFs is provided in Table 2 and the correlation statistics in Supplementary Table S1. Of note, five RFs (`original_firstorder_Energy, original_firstorder_10Percentile, original_firstorder_90Percentile, original_firstorder_Mean, original_ngtdm_Complexity`) were consistently significant across the three RF sets (Supplementary Figure S3). The scatter plots (Figure 4) depict the correlations between a representative radiomics feature (“`original_firstorder_Energy`”) and the three liver function indices.

**Figure 3.**
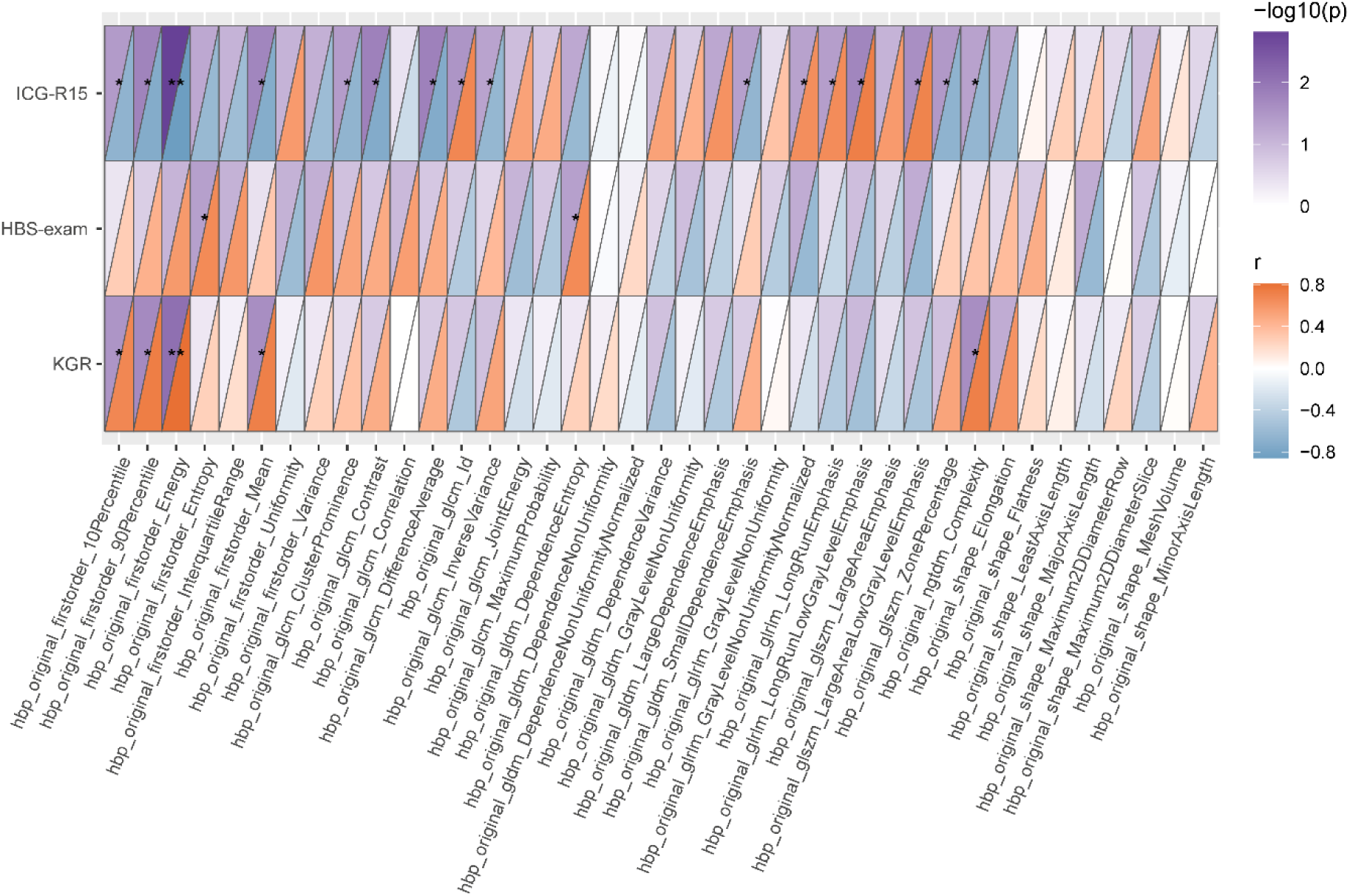
Correlation between radiomics features of hepatobiliary phase liver function indices. For each grid, the upper triangle represents p value (-log10(p)) and the lower triangle represents the correlation coefficient (r). * indicates p < 0.05, ** indicates p < 0.01.

**Figure 4.**
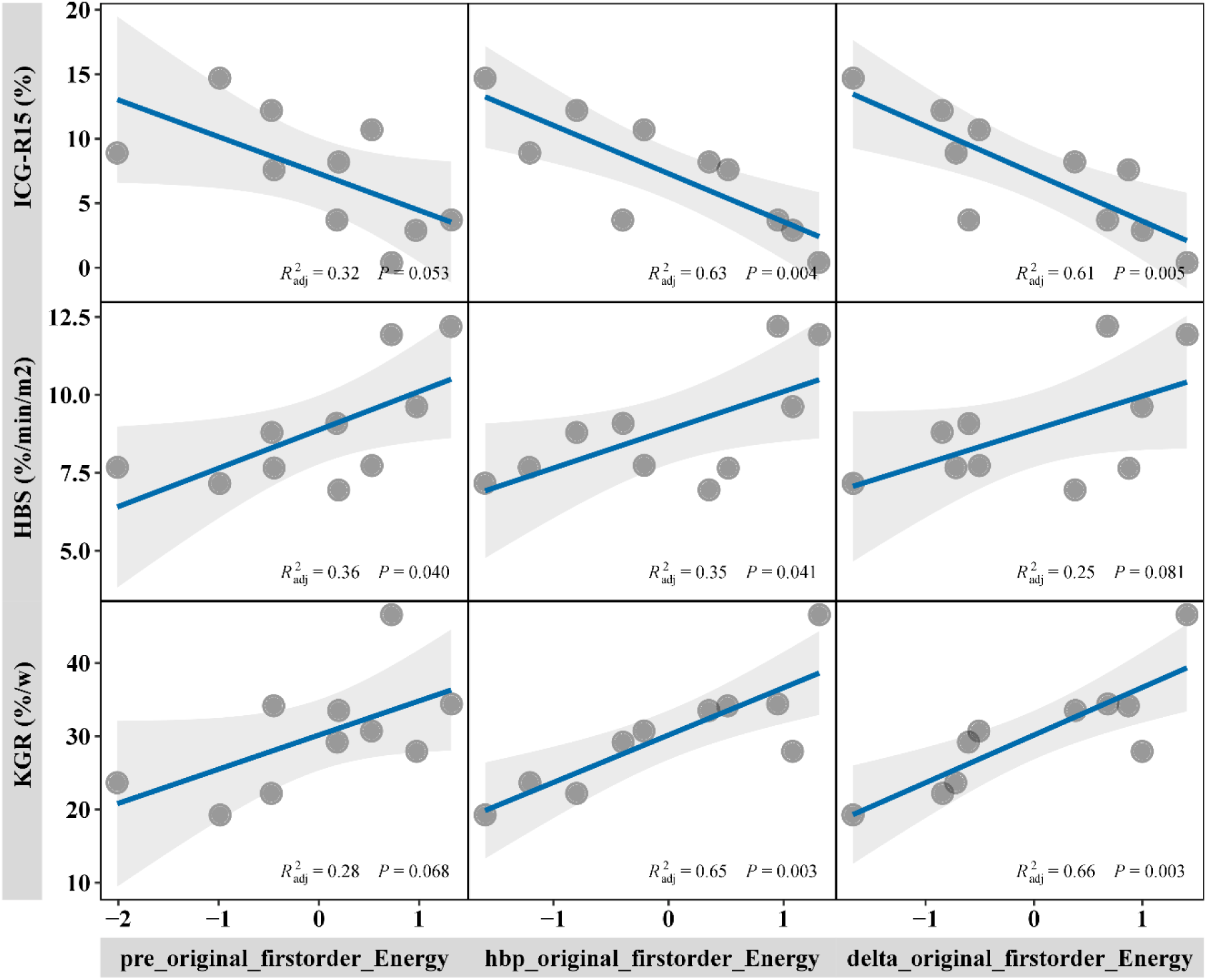
Correlation analysis of `original_firstorder_Energy` and liver function indices. Note that the radiomics feature was Z-score standardized when performing correlation analysis.

**Table 2.** Radiomics features with significant association with liver function indices.

| Pre-RF | HBP-RF | delta-RF |
| --- | --- | --- |
| original_firstorder_Energy | original_firstorder_Energy | original_firstorder_Energy |
| original_firstorder_10Percentile | original_firstorder_10Percentile | original_firstorder_10Percentile |
| original_firstorder_90Percentile | original_firstorder_90Percentile | original_firstorder_90Percentile |
| original_firstorder_Mean | original_firstorder_Mean | original_firstorder_Mean |
| original_ngtdm_Complexity | original_ngtdm_Complexity | original_ngtdm_Complexity |
| original_glcm_DifferenceEntropy | original_firstorder_Entropy | original_firstorder_Entropy |
| original_shape_MinorAxisLength | original_glcm_ClusterProminence | original_glcm_ClusterProminence |
|  | original_glcm_DifferenceAverage | original_glcm_DifferenceAverage |
|  | original_glrlm_GrayLevelNonUniformityNormalized | original_glrlm_GrayLevelNonUniformityNormalized |
|  | original_glszm_LargeAreaLowGrayLevelEmphasis | original_glcm_DifferenceEntropy |
|  | original_glszm_ZonePercentage | original_firstorder_InterquartileRange |
|  | original_gldm_SmallDependenceEmphasis | original_firstorder_MeanAbsoluteDeviation |
|  | original_glcm_Contrast | original_firstorder_Variance |
|  | original_glcm_Id | original_gldm_DependenceNonUniformity |
|  | original_glcm_InverseVariance | original_gldm_LargeDependenceEmphasis |
|  | original_glrlm_LongRunEmphasis | original_glcm_Idm |
|  | original_glrlm_LongRunLowGrayLevelEmphasis | original_glcm_JointEnergy |
|  | original_gldm_DependenceEntropy | original_glrlm_GrayLevelNonUniformity |
|  |  | original_glrlm_RunPercentage |
|  |  | original_firstorder_Uniformity |

*Note: Pre-RF and HBP-RF indicate the radiomics feature derived from gadoteric acid-enhanced MRI before contrast media injection and hepatobiliary phase. Radiomics features with green color indicate robustness across the three datasets, while those with blue color are robust across two datasets. Detailed definitions of radiomics features can be found at https://pyradiomics.readthedocs.io/en/latest/features.html.*

## Discussion

This study identified reproducible and repeatable radiomics features derived from gadoxetic acid-enhanced MRI that significantly correlated with liver function (represented by the ICG-R15 test, HBS, and KGR of the future liver remnant) in CRLM patients. By leveraging advanced imaging techniques and quantitative analysis methods, our work established the foundation of radiomics as a non-invasive tool in the quantitative liver function assessment.

Radiomics is increasingly recognized as a transformative tool for extracting quantitative imaging biomarkers that reflect underlying tissue characteristics [19, 20]. In this study, 214 RFs were extracted from gadoxetic acid-enhanced MRI pre-contrast media injection and HBP images. These RFs encompassed diverse categories such as shape, first-order statistics, and texture-based matrices. The reproducibility and repeatability of RFs were rigorously evaluated using inter-and intraclass correlation analyses, ensuring that the selected features were robust and consistent across delineation and processing conditions [18].

The study’s primary achievement lies in the identification of RFs, which correlated significantly with established liver function metrics. Specifically, 7 Pre-RFs demonstrated significant associations with liver function indices, reflecting the baseline liver parenchymal properties. Eighteen features were significantly associated with functional metrics, which highlighted the added value of hepatocyte-specific contrast enhancement in capturing liver function traits. Twenty delta-RFs were identified, which reflected the dynamic changes in liver parenchyma captured between unenhanced and HBP phased images. Among these, RFs such as `original_firstorder_Energy` were consistent across the three RF sets, suggesting their potential as universal biomarkers.

The correlation between gadoxetic acid-enhanced MRI and liver function metrics has been demonstrated in earlier research, primarily through simplified measures like liver-to-muscle signal intensity ratios [9, 21]. This study builds on such findings by expanding the scope to include radiomics features, which provide a high-dimensional and comprehensive representation of liver parenchymal properties. There are molecular foundations underpinning the correlation between radiomics features derived from gadoxetic acid-enhanced MRI, especially the hepatobiliary phase, and liver function metrics [5]. Gadoxetic acid uptake into hepatocytes is mediated primarily by organic anion-transporting polypeptides (OATPs) transporters, particularly OATP1B1 and OATP1B3, which are essential for hepatic function and bile production [5, 22]. Similarly, the ICG test and HBS exam rely on the hepatocyte’s ability to uptake and process specific substrates, both of which are also mediated by OATP activity [4, 23]. Therefore, radiomics features reflecting gadoxetic acid distribution inherently capture OATP-related liver functionality.

Traditional indices of gadoxetic acid-enhanced MRI evaluate signal intensity changes before contrast media injection and hepatobiliary phase, such as the liver relative enhancement [24, 25]. A large body of studies has applied delta radiomics in a variety of oncologic diseases [26, 27], but to our knowledge, not on liver-specific MRI. Delta-RFs capture dynamic changes of radiomics features between different MRI phases, adding a novel dimension to radiomics analysis. These features may offer unique insights into the liver’s functional reserve and regenerative potential. Interestingly, delta-RF had the largest number of repeatable and reproducible features, greater than Pre-RF and HBP-RF (20 vs 7, 18, respectively), to be significantly correlated with liver function metrics. This implies that dynamic changes of RF might be more informative and robust than RF derived from single-phased images.

Currently reproducibility and repeatability assessment of radiomics features remains essential in radiomics research, given that the manual segmentation of non-tumoral liver parenchyma is prone to operator variability. Still, manual segmentation is regarded as the ‘reference standard’[28]. The study’s focus on reproducibility through inter-and intraclass correlation analyses is a marked improvement over prior studies, which often lack detailed assessments of feature stability [12, 13]. The use of standardized protocols and software such as PyRadiomics and the Image Biomarker Standardization Initiative highlights the importance of methodological rigor in ensuring the reliability of extracted RFs. This step also sets a benchmark for future studies aiming to validate and expand upon the findings presented here.

Our findings have potential clinical implications. 1) The identified RFs could complement traditional metrics such as the ICG test and HBS to offer a more nuanced assessment of liver function and liver growth potential. By integrating imaging biomarkers with traditional clinical metrics, it contributes to comprehensively assess liver function for safer and more effective surgical interventions[29]. 2) One unique advantage of gadoxetic acid-enhanced MRI in evaluation of liver function lies in its potential to provide regional liver function information [30]. In this context, radiomics analysis can guide decisions regarding resection extent, risk stratification of post-hepatectomy liver failure, the necessity of preoperative portal vein embolization, or associating liver partition and portal vein ligation for staged hepatectomy. 3) The ability to assess liver function and growth capacity non-invasively using gadoxetic acid-enhanced MRI reduces the reliance on invasive tests, improving patient comfort and compliance. Importantly, radiomics analysis is based on often already available clinical MRI exams.

While the study provides valuable insights, several limitations warrant consideration. First, the study’s sample size of 10 patients from a single institution was a noticeable limitation, which might undermine the statistical power and generalizability of the findings. Future studies should aim to validate these results in larger, multicenter cohorts[31]. Second, this study only focused on radiomics features derived from the raw intensity images and did not evaluate different filters (for instance, wavelet) transformed features. Future studies should incorporate these filters to expand the radiomics feature set, evaluate their added value, and validate their utility in larger, diverse cohorts. Third, the study subjects were limited to patients with CRLM. Whether the findings can be extrapolated to patients with primary liver cancers such as hepatocellular carcinoma or non-cancer settings e.g. non-alcoholic fatty liver disease requires further investigation. Last, despite the rigorous reproducibility assessment, manual segmentation of the liver parenchyma remains a time-intensive process with potential for inter-operator variability. Automated segmentation by deep learning techniques should be explored in future research to improve efficiency and reduce variability.

In conclusion, a set of reliable and clinically relevant radiomics features was identified from preoperative gadoxetic acid-enhanced MRI, which provides a foundation for future research targeting non-invasive quantitative liver function assessment and post-hepatectomy liver failure prediction. Our findings highlight the potential of gadoxetic acid-enhanced MRI-based radiomics features as a liver function biomarker and underscore the promise of radiomics in advancing personalized medicine.

## Ethics approval and consent to participate

The research protocol was approved by the Regional Ethical Review Board (Dnr 2012/583-31/4) and informed consent was obtained from all participating patients.

## Trial registration

This study is a secondary analysis of a prospective cohort that has been registered at https://clinicaltrials.gov/ with an identifier NCT03140917.

## Consent for publication

Not applicable.

## Availability of data and materials

The original contributions presented in the study are included in the article/ Supplementary Materials. Further inquiries can be directed to the corresponding author.

## Data Availability

The original contributions presented in this study are included in the article/supplementary material. Further inquiries can be directed to the corresponding author.

## Acknowledgements

No.

## Supplementary materials

Figure S1. Heatmap of the correlation coefficients of the radiomics feature from pre-contrast media injection images (A), the hepatobiliary phase (B), and the delta RF (C).

Figure S2. Correlations between radiomics features of pre-contrast media injection images (A) and the delta-RF (B) and liver function indices.

Figure S3. Venn diagram shows the overlapping of different radiomics feature sets.

Table S1. List of radiomics features with detailed information on the correlation statistic with liver function indices

## List of abbreviations

CRLM: Colorectal liver metastasis
HBP: Hepatobiliary phase
HBS: Hepatobiliary scintigraphy
ICG: Indocyanine green
ICG-R15: ICG retention rate at 15 minutes
KGR: Kinetic growth rate
MRI: Magnetic resonance imaging
OATPs: Organic anion-transporting polypeptides
RF: Radiomics feature.

