## Supplemental Figures and Tables for "Liver functional radiomics of gadoxetic acid-enhanced magnetic resonance imaging: A proof-of-concept study"

### Supplementary materials

Figure S1. Heatmap of the correlation coefficients of the radiomics feature from pre-contrast media injection images (A), the hepatobiliary phase (B), and the delta RF (C).

A

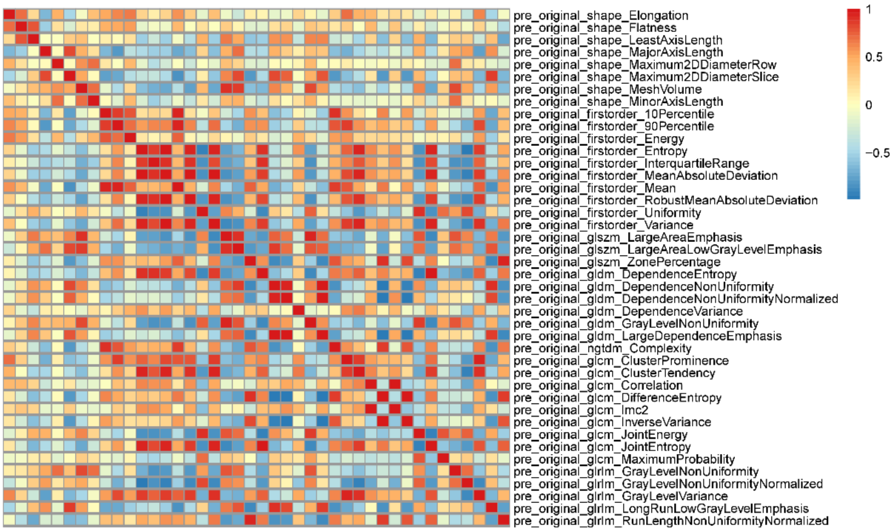


B

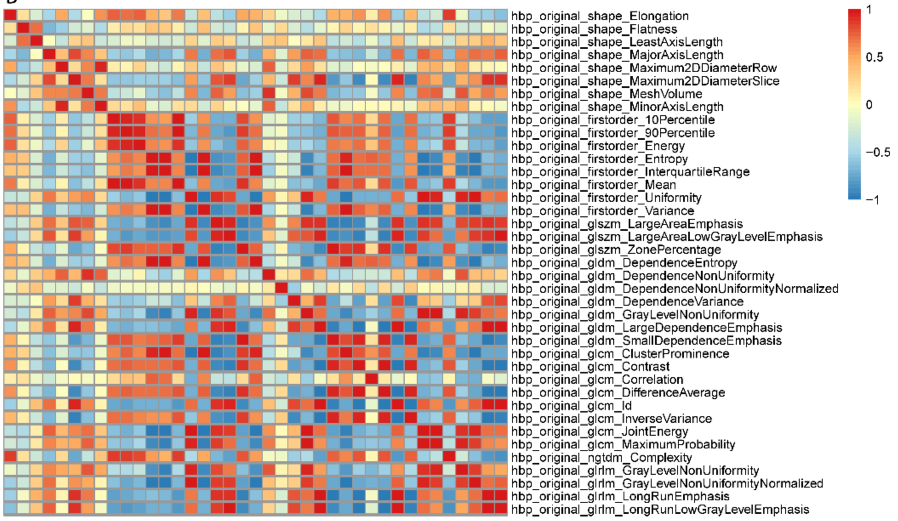


C

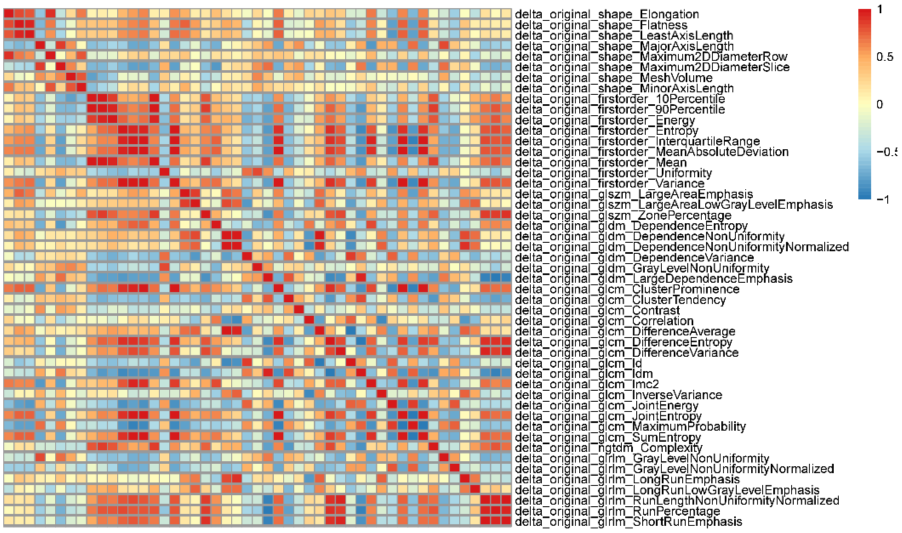

Figure S2. Correlations between radiomics features of pre-contrast media injection images (A) and the delta-RF (B) and liver function indices. For each grid, the upper triangle represents p value (-log10(p)) and the lower triangle represents the correlation coefficient (r). * indicates p < 0.05, ** indicates p < 0.01.

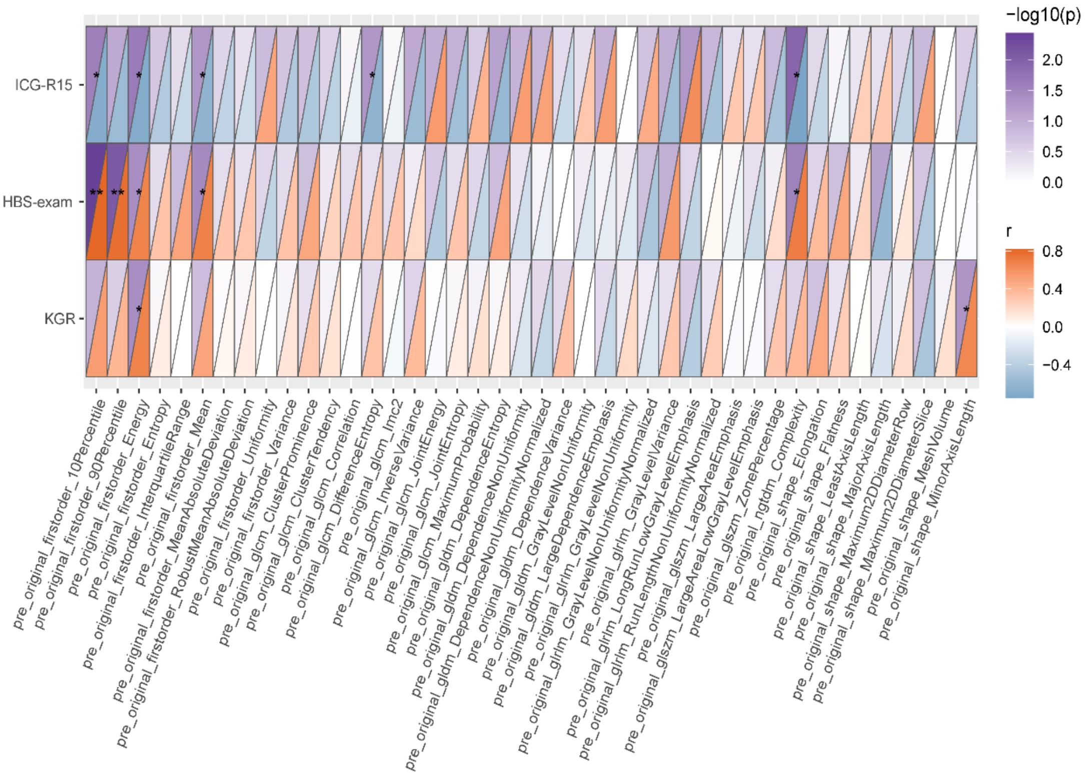


A

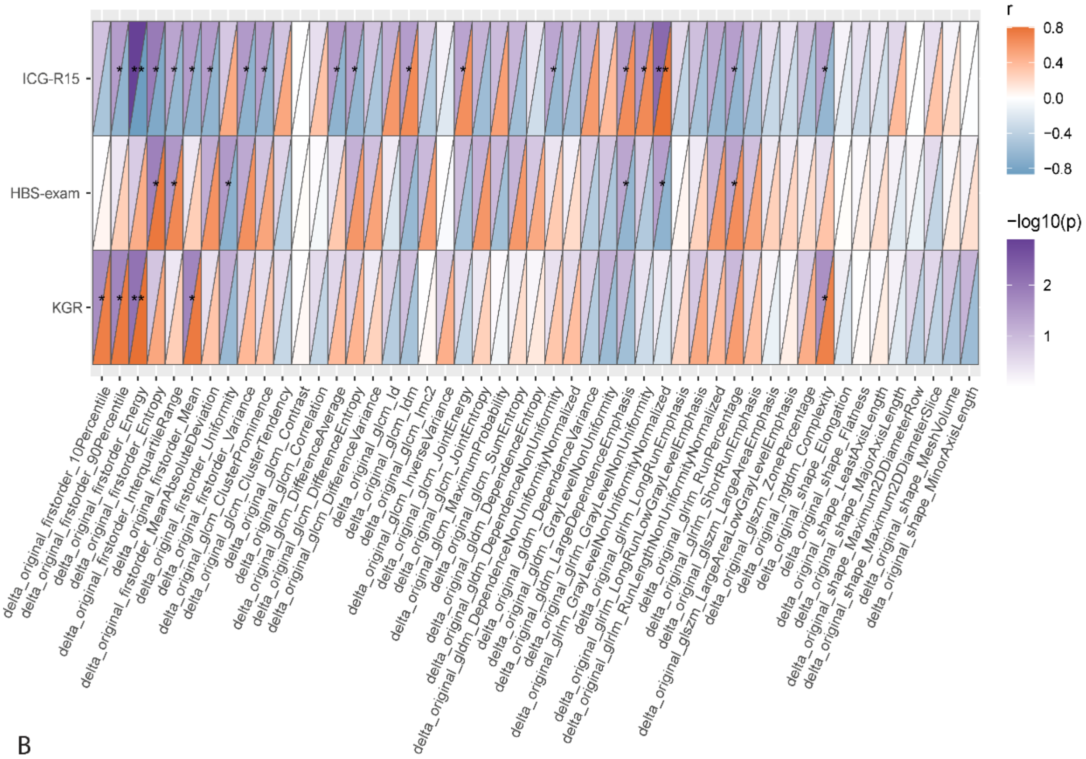


B

Figure S3. Venn diagram shows the overlapping of different radiomics feature sets. Five radiomics features demonstrate robust correlation with liver function metrics across the three radiomics feature sets.

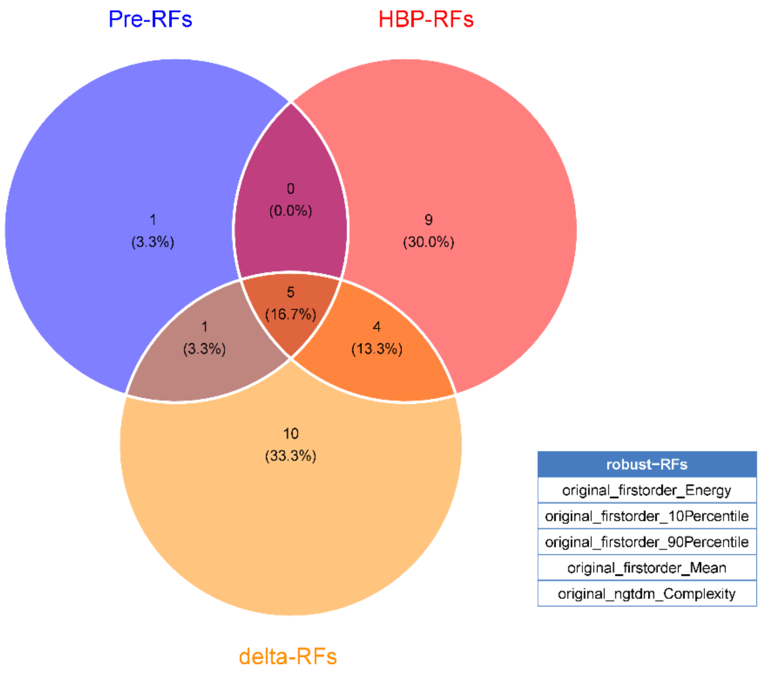

Table S1. List of radiomics features with detailed information on the correlation statistic with liver function indices

S1A. Pre-RFs

| Radiomics feature | Liver function/growth index | Correlation coefficient ® | p value |
|---|---|---|---|
| **original_firstorder_10Percentile** | ICG_R15 | -0.69326458 | 0.026215485 |
| **original_firstorder_Energy** | ICG_R15 | -0.705170431 | 0.022737737 |
| **original_firstorder_Mean** | ICG_R15 | -0.632221765 | 0.0498461 |
| **original_ngtdm_Complexity** | ICG_R15 | -0.753802874 | 0.011794786 |
| **original_glcm_DifferenceEntropy** | ICG_R15 | -0.632221765 | 0.0498461 |
| **original_firstorder_10Percentile** | HBS_test | 0.819606358 | 0.00370353 |
| **original_firstorder_90Percentile** | HBS_test | 0.778119096 | 0.008033138 |
| **original_firstorder_Energy** | HBS_test | 0.672727273 | 0.039381409 |
| **original_firstorder_Mean** | HBS_test | 0.684848485 | 0.035091538 |
| **original_ngtdm_Complexity** | HBS_test | 0.709090909 | 0.027514119 |
| **original_shape_MinorAxisLength** | KGR | 0.648484848 | 0.049042632 |
| **original_firstorder_Energy** | KGR | 0.672727273 | 0.039381409 |

S1B. HBP-RFs

| Radiomics feature | Liver function/growth index | Correlation coefficient ® | p value |
|---|---|---|---|
| **original_firstorder_10Percentile** | ICG_R15 | -0.661585366 | 0.037206185 |
| **original_firstorder_90Percentile** | ICG_R15 | -0.729486652 | 0.01664688 |
| **original_firstorder_Energy** | ICG_R15 | -0.857146817 | 0.00152784 |
| **original_firstorder_Mean** | ICG_R15 | -0.723407597 | 0.018047748 |
| **original_glszm_LargeAreaLowGrayLevelEmphasis** | ICG_R15 | 0.699091375 | 0.024470991 |
| **original_glszm_ZonePercentage** | ICG_R15 | -0.668696098 | 0.034509542 |
| **original_gldm_SmallDependenceEmphasis** | ICG_R15 | -0.644379876 | 0.044311514 |
| **original_glcm_ClusterProminence** | ICG_R15 | -0.656537987 | 0.039204386 |
| **original_glcm_Contrast** | ICG_R15 | -0.735565708 | 0.015323456 |
| **original_glcm_DifferenceAverage** | ICG_R15 | -0.735565708 | 0.015323456 |
| **original_glcm_Id** | ICG_R15 | 0.680854209 | 0.030211171 |
| **original_glcm_InverseVariance** | ICG_R15 | -0.632221765 | 0.0498461 |
| **original_ngtdm_Complexity** | ICG_R15 | -0.638300821 | 0.047024453 |
| **original_glrlm_GrayLevelNonUniformityNormalized** | ICG_R15 | 0.638300821 | 0.047024453 |
| **original_glrlm_LongRunEmphasis** | ICG_R15 | 0.650458932 | 0.041705451 |
| **original_glrlm_LongRunLowGrayLevelEmphasis** | ICG_R15 | 0.723407597 | 0.018047748 |
| **original_firstorder_Entropy** | HBS_test | 0.660606061 | 0.044026553 |
| **original_gldm_DependenceEntropy** | HBS_test | 0.660606061 | 0.044026553 |
| **original_firstorder_10Percentile** | KGR | 0.680854209 | 0.030211171 |

| | | | |
|---|---|---|---|
| **original_firstorder_90Percentile** | KGR | 0.733333333 | 0.021166481 |
| **original_firstorder_Energy** | KGR | 0.806060606 | 0.008235571 |
| **original_firstorder_Mean** | KGR | 0.721212121 | 0.024194588 |
| **original_ngtdm_Complexity** | KGR | 0.721212121 | 0.024194588 |

S1C. delta-RFs

| Radiomics feature | Liver function/growth index | Correlation coefficient ® | p value |
|---|---|---|---|
| **original_firstorder_90Percentile** | ICG_R15 | -0.66261704 | 0.036806402 |
| **original_firstorder_Energy** | ICG_R15 | -0.86322587 | 0.001293815 |
| **original_firstorder_Entropy** | ICG_R15 | -0.75988193 | 0.010758005 |
| **original_firstorder_InterquartileRange** | ICG_R15 | -0.63719512 | 0.047529531 |
| **original_firstorder_MeanAbsoluteDeviation** | ICG_R15 | -0.66261704 | 0.036806402 |
| **original_firstorder_Mean** | ICG_R15 | -0.66261704 | 0.036806402 |
| **original_firstorder_Variance** | ICG_R15 | -0.67477515 | 0.032311809 |
| **original_gldm_DependenceNonUniformity** | ICG_R15 | -0.63222177 | 0.0498461 |
| **original_gldm_LargeDependenceEmphasis** | ICG_R15 | 0.656537987 | 0.039204386 |
| **original_glcm_ClusterProminence** | ICG_R15 | -0.65653799 | 0.039204386 |
| **original_glcm_DifferenceAverage** | ICG_R15 | -0.6686961 | 0.034509542 |
| **original_glcm_DifferenceEntropy** | ICG_R15 | -0.65045893 | 0.041705451 |
| **original_glcm_Idm** | ICG_R15 | 0.662617043 | 0.036806402 |
| **original_glcm_JointEnergy** | ICG_R15 | 0.638300821 | 0.047024453 |
| **original_ngtdm_Complexity** | ICG_R15 | -0.63830082 | 0.047024453 |
| **original_glrlm_GrayLevelNonUniformity** | ICG_R15 | 0.668696098 | 0.034509542 |
| **original_glrlm_GrayLevelNonUniformityNormalized** | ICG_R15 | 0.796356262 | 0.005836997 |
| **original_glrlm_RunPercentage** | ICG_R15 | -0.65653799 | 0.039204386 |
| **original_firstorder_Entropy** | HBS_test | 0.76969697 | 0.013671782 |
| **original_firstorder_InterquartileRange** | HBS_test | 0.680854209 | 0.030211171 |
| **original_firstorder_Uniformity** | HBS_test | -0.6969697 | 0.031141095 |
| **original_gldm_LargeDependenceEmphasis** | HBS_test | -0.64848485 | 0.049042632 |
| **original_glrlm_GrayLevelNonUniformityNormalized** | HBS_test | -0.67272727 | 0.039381409 |
| **original_glrlm_RunPercentage** | HBS_test | 0.648484848 | 0.049042632 |
| **original_firstorder_10Percentile** | KGR | 0.733333333 | 0.021166481 |
| **original_firstorder_90Percentile** | KGR | 0.757575758 | 0.015920829 |
| **original_firstorder_Energy** | KGR | 0.806060606 | 0.008235571 |
| **original_firstorder_Mean** | KGR | 0.757575758 | 0.015920829 |
| **original_ngtdm_Complexity** | KGR | 0.721212121 | 0.024194588 |
